# Distinct rates of VUS reclassification are observed when subclassifying VUS by evidence level

**DOI:** 10.1101/2024.11.13.24317242

**Authors:** Gwendolyn Bennett, Izabela Karbassi, Wenjie Chen, Steven M. Harrison, Matthew S. Lebo, Linyan Meng, Narasimhan Nagan, Robert Rigobello, Heidi L. Rehm

## Abstract

**Purpose:** Genetic testing commonly yields a plethora of variants of uncertain significance (VUS) that can lead to ongoing uncertainty for patients and their caregivers. While all VUS hold uncertainty, some VUS have more evidence in support of pathogenicity while others have more evidence of a benign role. Sharing these nuances can help guide the investment in follow-up clinical and research investigations and may, at times, influence medical decision-making despite appreciated uncertainty.

**Methods:** Four clinical laboratories have been subclassifying VUS to help prioritize investigation and guide reporting decisions. Each laboratory developed a distinct approach for how these subclasses are used in their laboratories and, in some cases, displayed on reports. We examined the composition of each laboratory’s VUS subclasses and the likelihood variants from each subclass were reclassified towards pathogenic or benign.

**Results:** We found that variants in the lowest subclass of VUS were never reclassified as likely pathogenic (LP) or pathogenic (P), while those in the highest subclass were much more likely to be reclassified as P/LP.

**Conclusion:** Given that forthcoming professional guidance in variant classification will advise the use of VUS subclasses, the experience of our laboratories in using VUS subclasses can inform future practices.

## Introduction

The rapid advancement of genetic testing has revolutionized our understanding of the genetic basis of many diseases and conditions. However, the interpretation of genetic test results remains a challenging practice, particularly when many reports contain variants of uncertain significance (VUS).^1^ These genetic variants, without a clear association with disease, do not meet pathogenic or benign as defined by current professional guidelines for the classification of sequence variants published by the American College of Genetics and Genomics (ACMG) and Association for Molecular Pathology (AMP) in 2015.^2^ The 2015 ACMG/AMP guidelines, and subsequent clarifications from the Clinical Genome Resource,^3,4^ indicate the use of “likely benign” and “likely pathogenic” should signify a 90-99% confidence of a particular variant being benign or pathogenic. This leaves the VUS category spanning the intervening 80% range of confidence of pathogenicity (10% - 90%). To help in the classification of sequence variants that fall into this expansive VUS category, some laboratories and other professional organizations developed independent criteria to divide this category into subclasses.^5–8^ Of note, subclasses for the VUS category were recognized by the 2015 ACMG/AMP guidelines as allowable, but with limited guidance as to how to define, utilize, and report them.

Here, we present the experience of four laboratories who have been applying VUS subclasses for 10 years (Baylor Genetics), 13 years (Labcorp), 15 years (Mass General Brigham Laboratory for Molecular Medicine (MGB LMM)), and 14 years (Quest Diagnostics). While each laboratory developed their own criteria for subdividing the VUS category, all four created three subclasses which are similar in concept and evidence strengths to the system proposed for the next version of the 2015 ACMG/AMP guidelines: VUS-high, VUS-mid, and VUS-low (Figure 1). Variants classified into the VUS-high subclass have evidence to indicate the variant could be pathogenic, but insufficient to classify the variant as likely pathogenic. On the other end, variants in the VUS-low subclass have evidence to suggest the variant may be benign, but still insufficient to classify the variant as likely benign. Variants in the VUS-mid subclass are more equivocal and may have equal but conflicting evidence regarding pathogenicity, or no evidence at all.

**Figure 1.**
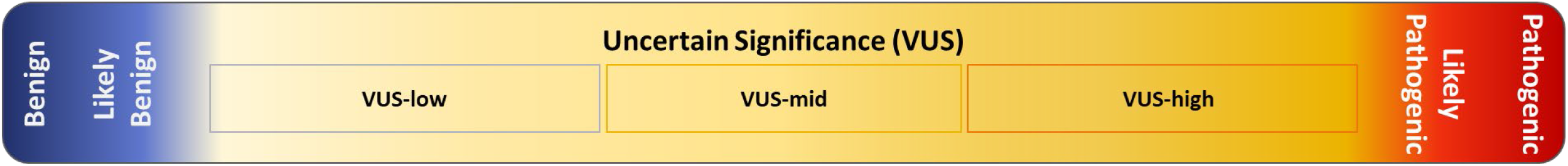
Variant pathogenicity classification terminology. Sequence variation is typically assigned to one of five pathogenicity classifications based on current professional guidelines. At left, in blue, are the classifications benign and likely benign. At right, in red, are classifications for likely pathogenic and pathogenic. Variants of uncertain significance (VUS) are shown in yellow. Some laboratories divide VUS into three subclasses, which are similar in concept and evidence strengths to the system proposed for the next version of the professional sequence variant classification standards: VUS-low, VUS-mid, and VUS-high. Variants within these subclasses have graded levels of evidence that is either insufficient to reach likely pathogenic or likely benign, or is conflicting.

For each of the four laboratories, the use of VUS subclasses has been driven by clinician and internal laboratorian feedback – both wanting guidance on prioritizing follow-up efforts to investigate VUS, as well as reducing the negative impact to medical practices. Each laboratory independently developed and implemented VUS subclassification rubrics for internal management of variant classifications, as well as aiding in conversations regarding variant reclassification or participation in VUS clarification/resolution programs, where available. The approaches to how these VUS subclasses were presented externally, either on patient reports or in data sharing efforts, varied amongst the laboratories. Baylor Genetics uses the subclass to determine inclusion of the variant on patient reports for hereditary cancer panels. VUS-low variants are in general not reported in clinical diagnostic or screening tests, while VUS-high and VUS-mid are included on reports as a single VUS category. However, the subclass is not specified on patient reports. Labcorp combines VUS subclasses into a single VUS category for reporting carrier screening. The VUS subclasses are reported with specific wording in some but not all diagnostic settings, notably for hereditary cancer tests. Detailed evidence summaries supporting the underlying subclass are submitted to ClinVar and made available upon client/provider request. MGB LMM includes wording specific to the subclass in the evidence summary section of all patient reports where VUS are reported (i.e., symptomatic testing) and includes VUS-high on reports for certain clinical research studies where VUS are typically not reported (e.g., healthy population screening). Quest Diagnostics provides wording specific to, and a visual representation of, the variant classification and VUS subclasses on clinical reports from Athena Diagnostics. Providing subclass language on all sequencing tests that report VUS is being implemented; at the time of writing, VUS subclasses are combined for patient reporting in other Quest tests as well as for ClinVar submissions. More details specific to each laboratory’s implementations can be found in Table S1.

In a recent publication in which Quest Diagnostics and MGB LMM participated,^1^ it was demonstrated that no VUS-low variants were reported in any clinical exome or genome sequencing (ES/GS) results, where professional decision-making informs which VUS are reported. This was in contrast to multi-gene panel (MGP) reports where 22% of VUS were of the VUS-low category, given typical MGP reporting practices of including all VUS regardless of evidence level or likelihood of causality. While this MGP reporting strategy made sense when panel sizes were small and patient indications largely matched the phenotypes associated to the included genes, the current era of very large MGPs has lessened the gene-to-phenotype match and increased the number of variants included on reports. De-emphasizing VUS unlikely to be causal in MGP testing is challenging given the difficulty laboratories have gaining access to phenotype to allow application of phenotype-specificity evidence (PP4 in the 2015 ACMG/AMP guidelines), which may convert a VUS-low to a higher classification. In contrast, ES/GS requires the provision of phenotype data, allowing this information to guide variant classification and case interpretation. Even with these common practices in use, limited guidance exists from professional organizations around VUS reporting except for carrier screening.^9,10^

Here, we investigated the directionality and odds of reclassification for variants across our four laboratories with a focus on the VUS subclasses. We have identified distinct trends that are meaningful for clinical laboratories, healthcare providers, and patients in addressing the challenges surrounding VUS as well as informing policy development in clinical genetic test reporting.

## Materials and Methods

We collected variant classification data from each laboratory during an overall study period from January 2016 to April 2023, though exact periods varied by laboratory as indicated below. For each of 151,368 variants, their earliest classification during the study period and their current classification were captured. Classification criteria were based on 2015 ACMG/AMP guidelines, in conjunction with additional ClinGen guidance^2,11^ and laboratory-specific criteria. Copy number variants (CNVs) were not included. During the period of the study, bioinformatics pipelines, classification criteria, and other steps within the variant curation process did not significantly change for any of the four laboratories. The number of variants where the classification either moved up towards a pathogenic classification, moved down towards a benign classification, or remained unchanged were determined. Variants that were evaluated only once during the study period were treated as unchanged. Additionally, Quest Diagnostics and Labcorp were able to track classification history for variants that were evaluated multiple times during the study period. Note, variants or variant classification events do not correlate with the number of specimens tested.

For each laboratory, variants and variant classification events were identified as follows:

### Baylor Genetics

A total of 40,828 variants identified as part of hereditary cancer testing performed from June 2020 to April 2023 were compiled. For each of these variants, their classification when first identified by testing and their current classification were captured. Variants predicted by the bioinformatics pipeline as benign or likely benign based on population frequency were not independently curated and so were not captured in this dataset.

### Labcorp

A total of 28,411 variants with classification events between January 2022 and February 2024 were compiled. This dataset spans variants identified as part of diagnostic panels for a variety of indications including but not limited to cardiac, cancer, neurology, and metabolism and as part of diagnostic exome evaluations. Additionally, variants were also classified within genes comprising expanded carrier screening panels for recessive disorders. For each variant, all classification events occurring during the study period were collected.

### MGB LMM

A total of 44,113 variants classified or reclassified for Mendelian disorders from January 2016 to April 2023 were compiled. MGB LMM’s classifications include a higher volume of inherited cardiovascular disorders, sensorineural hearing loss, and variants in the ACMG Secondary Findings genes.^12^ Variants were included if the first classification or one or more reassessments happened during the study period. Dates of initial classification prior to April 2016 were captured on variants reclassified during the study period.

### Quest Diagnostics

A total of 38,016 variants with classification events from January 2017 to March 2023 were compiled. These variants were identified during testing performed for neurological, endocrine, nephrotic, hereditary cancer and other inherited genetic disorders, or evaluated as needed for other reasons. For each variant, all classification events occurring during the study period were collected.

### Odds ratios

For each VUS subclass, the odds of a variant moving up (towards pathogenic) was compared to the odds of a variant moving up if no VUS subclasses were used. Calculations were performed using MedCalc’s odds ratio calculator.^13^

## Results

The distribution of variants from four laboratories and across six variant classifications from likely benign to pathogenic, including the three VUS subclasses, is shown in Figure 2A and Table 1. Variants classified as benign at the time of data gathering were excluded from Figure 2 given that each laboratory managed this class differently, with some filtering out high frequency variants without formal classification. The largest fraction was made up of variants classified as likely benign (LB, 36.2%), while the smallest was the VUS-high subclass (4.0%). While each laboratory contributed a similar share of the total variants, the distribution across variant classifications was unique for each laboratory (Figure S1). Differences are likely due to a combination of factors, such as evidence thresholds for classification, the time between initial testing and any subsequent review, the timeframe for this study, distinct sets of genes tested in each lab, and attributes of those genes that contribute to disease.

**Table 1.**
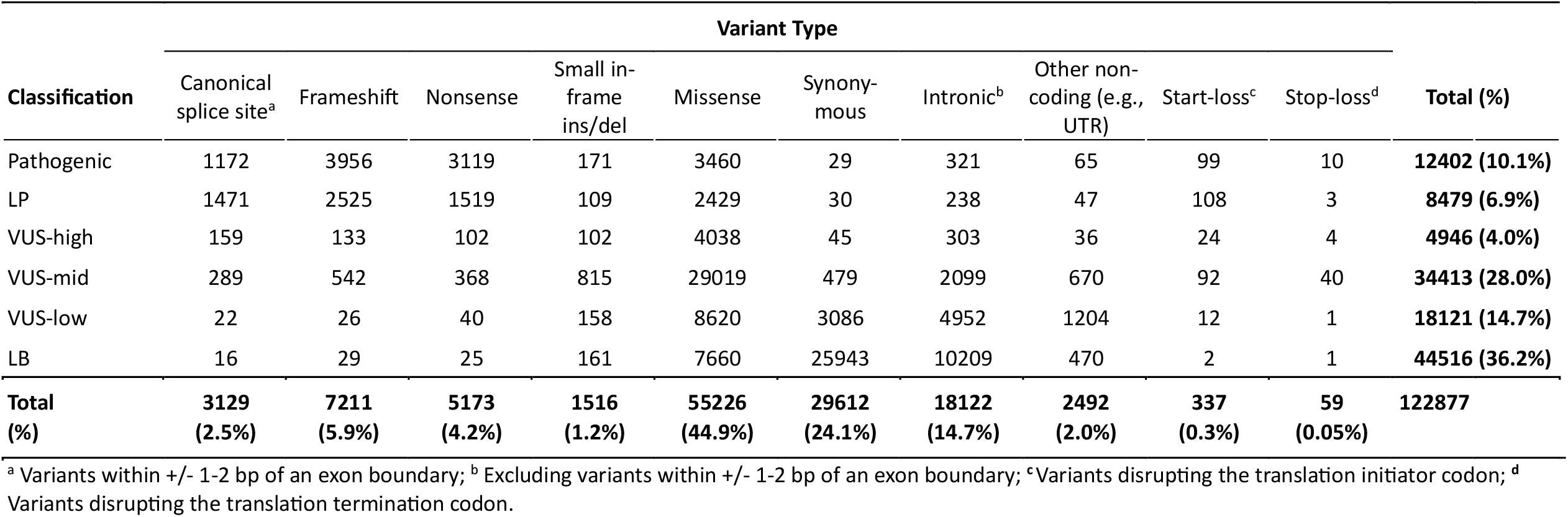
Distribution of variants across six pathogenicity classes from likely benign to pathogenic, including three VUS subclasses, grouped by variant type. Data from four contributing laboratories.

| Classification | Variant Type |  |  |  |  |  |  |  |  |  | Total (%) |
| --- | --- | --- | --- | --- | --- | --- | --- | --- | --- | --- | --- |
|  | Canonical splice site<sup>a</sup> | Frameshift | Nonsense | Small in-frame ins/del | Missense | Synonymous | Intronic<sup>b</sup> | Other non-coding (e.g., UTR) | Start-loss<sup>c</sup> | Stop-loss<sup>d</sup> |  |
| Pathogenic | 1172 | 3956 | 3119 | 171 | 3460 | 29 | 321 | 65 | 99 | 10 | 12402 (10.1%) |
| LP | 1471 | 2525 | 1519 | 109 | 2429 | 30 | 238 | 47 | 108 | 3 | 8479 (6.9%) |
| VUS-high | 159 | 133 | 102 | 102 | 4038 | 45 | 303 | 36 | 24 | 4 | 4946 (4.0%) |
| VUS-mid | 289 | 542 | 368 | 815 | 29019 | 479 | 2099 | 670 | 92 | 40 | 34413 (28.0%) |
| VUS-low | 22 | 26 | 40 | 158 | 8620 | 3086 | 4952 | 1204 | 12 | 1 | 18121 (14.7%) |
| LB | 16 | 29 | 25 | 161 | 7660 | 25943 | 10209 | 470 | 2 | 1 | 44516 (36.2%) |
| **Total (%)** | **3129 (2.5%)** | **7211 (5.9%)** | **5173 (4.2%)** | **1516 (1.2%)** | **55226 (44.9%)** | **29612 (24.1%)** | **18122 (14.7%)** | **2492 (2.0%)** | **337 (0.3%)** | **59 (0.05%)** | **122877** |

<sup>a</sup> Variants within +/- 1-2 bp of an exon boundary; <sup>b</sup> Excluding variants within +/- 1-2 bp of an exon boundary; <sup>c</sup> Variants disrupting the translation initiator codon; <sup>d</sup> Variants disrupting the translation termination codon.

**Figure 2.**
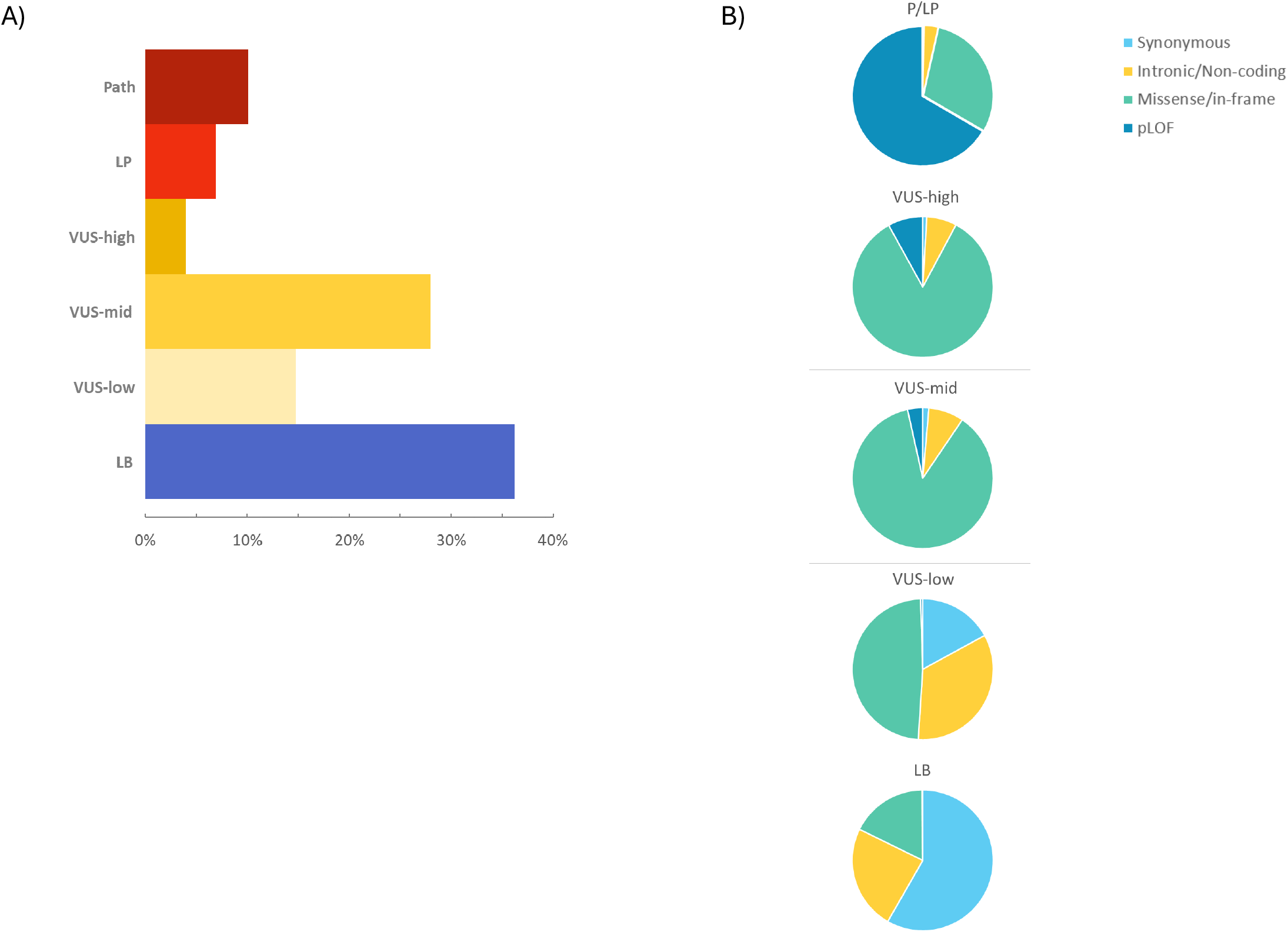
Variant classification distribution. (A) Distribution of 122,877 variants from four laboratories (Baylor Genetics, Labcorp, Mass General Brigham Laboratory for Molecular Medicine, Quest Diagnostics) across six variant classifications from likely benign to pathogenic, including three VUS subclasses. (B) Distribution of variant types within each variant classification. Variants disrupting the translation initiator codon (start-loss) and translation termination codon (stop-loss) are not included and represent < 0.4% of variants, combined (Table 1). Benign variants are not shown. P/LP, pathogenic or likely pathogenic; Path, pathogenic; LP, likely pathogenic; VUS; variant of uncertain significance; LB, likely benign; intronic, intronic variants excluding those within +/-1-2 bp of an exon boundary; non-coding, variants not directly altering a protein-coding region, excluding intronic variants; in-frame, small insertions or deletions altering a protein-coding region yet maintaining the translational reading frame.

We examined the types of variants that made up each classification (Figure 2B). Unsurprisingly, the vast majority (65.9%) of pathogenic and likely pathogenic (P/LP) variants were those that are predicted to lead to loss of function (pLOF): frameshift, nonsense, and canonical splice site variants. Across all six classifications, pLOF variants made up only 12.6% of all variants (Table 1). Each of the three VUS subclasses were primarily composed of missense and small in-frame deletion or insertion variants. However, the VUS-low subclass contained a substantially higher portion of synonymous (17.0%) and non-coding (intronic, UTR, etc.; 34.0%) variants compared to the VUS-mid (9.4%, combined) or VUS-high (7.8%, combined) subclasses.

Across the study period, we were able to track changes in variant classification and whether the classification moved up towards a pathogenic classification, moved down towards a benign classification, or remained unchanged. Overall, when variants changed classification or subclassification, they were 4.3 times more likely to move down towards benign, rather than up towards pathogenic (Table 2). As expected, variants that started with a classification of benign or pathogenic were the most stable, with less than 1% moving away from these starting classifications (Figure 2; n = 35, n = 87, respectively, Table 2). Variants with a starting classification of likely benign, VUS, or likely pathogenic were also reasonably stable, with only 2.1%, 2.6% and 3.3%, respectively, moving away from each of these starting classifications (Figure 3, Figure S2).

**Table 2.** Distribution of variants from four laboratories that changed classification “down” (towards benign) or “up” (towards pathogenic).

| Starting classification | Ending Classification |  |  |  |  |  |  | Total reclassified | Total moved down | Total moved up | Odds (Down : Up) | Odds ratio (95% CI) |
| --- | --- | --- | --- | --- | --- | --- | --- | --- | --- | --- | --- | --- |
|  | Benign | LB | VUS-low | VUS-mid | VUS-high | LP | Path |  |  |  |  |  |
| **Pathogenic** | 0 | 0 | 1 | 7 | 2 | 77 | - | **87** | 87 | - | - |  |
| **LP** | 0 | 3 | 2 | 18 | 39 | - | 222 | **284** | 62 | 222 | 1 : 3.6 |  |
| **VUS-high** | 0 | 2 | 35 | 265 | - | 182 | 35 | **519** | 302 | 217 | 1.4 : 1 | 2.90* (2.34 - 3.60) |
| **VUS-mid** | 100 | 305 | 1497 | - | 233 | 69 | 14 | **2218** | 1902 | 316 | 6.0 : 1 | 0.67* (0.56 - 0.80) |
| **VUS-low** | 234 | 570 | - | 83 | 6 | 0 | 0 | **893** | 804 | 89 | 9.0 : 1 | 0.45* (0.35 - 0.58) |
| *VUS (total)* | *334* | *877* | - | - | - | *251* | *49* | *1511<sup>b</sup>* | *1211* | *300* | *4.0 : 1* | *Ref. category* |
| **LB** | 868 | - | 40 | 14 | 0 | 0 | 0 | **922** | 868 | 54 | 16 : 1 |  |
| **Benign** | - | 32 | 3 | 0 | 0 | 0 | 0 | **35** | - | 35 | - |  |
| **Overall<sup>a</sup>** |  |  |  |  |  |  |  | **4958** | **4025** | **933** | **4.3 : 1** |  |

<sup>a</sup> Using 7 classifications, including 3 VUS subclasses; <sup>b</sup> Total VUS reclassified to non-VUS; \* p < 0.0001

**Figure 3.**
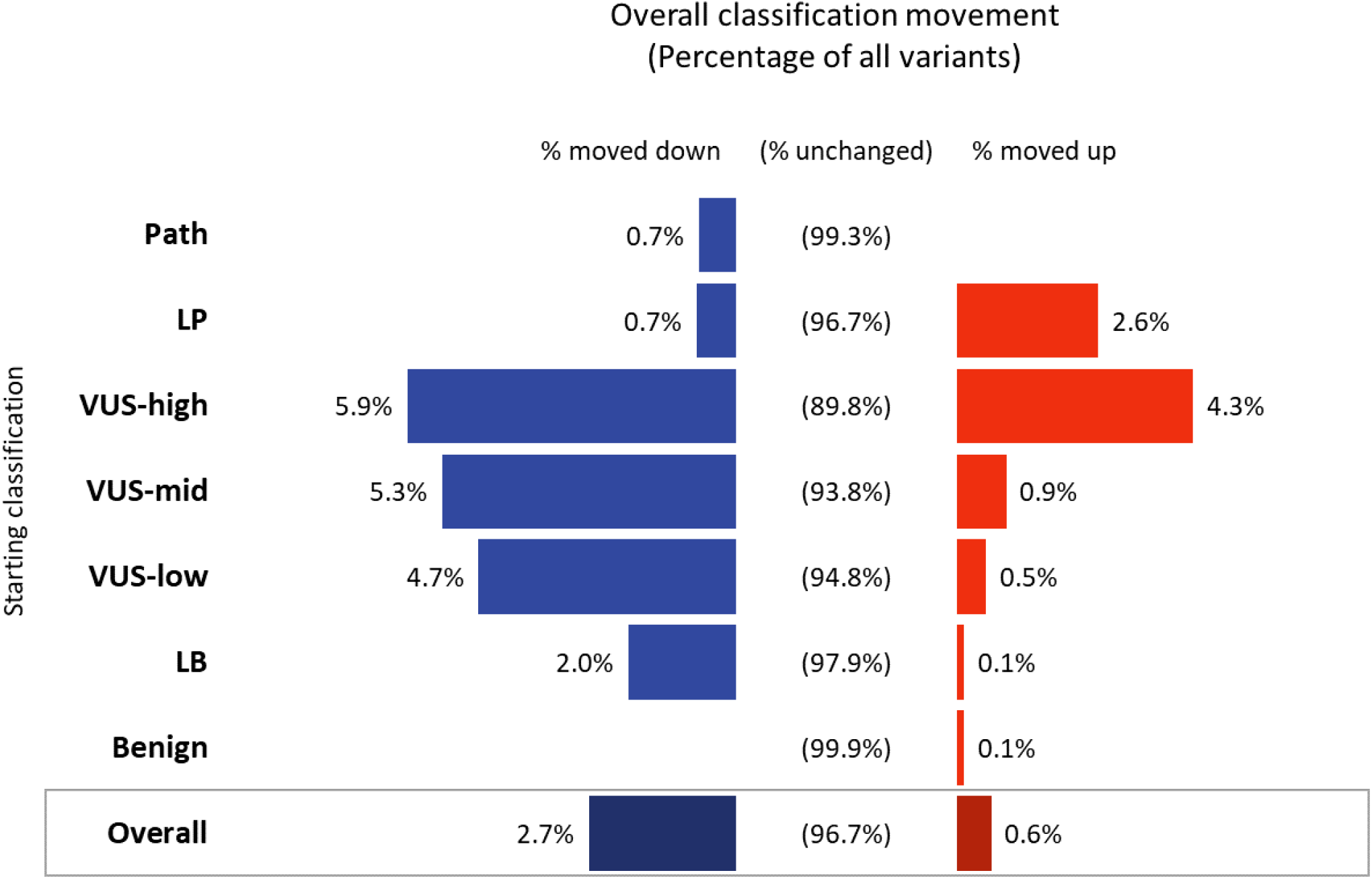
Classification movement by overall direction. The figure shows the percentage of variants aggregated across the four laboratories that did or did not change classification. Variants’ initial (starting) classification during the study period is shown at left. Blue bars, the percentage of variants whose classification moved at least one category “down” (toward benign). Red bars, the percentage of variants whose classification moved at least one category “up” (toward pathogenic). Variants that were not re-evaluated or whose classification did not change during the study period are shown as “unchanged.” Abbreviations as in Figure 2.

Among all categories, the VUS-high subclass had the highest percentage of variant reclassifications (10.2%, Figure 4), yet, since this category makes up only 4.0% of current classifications across the four labs (Figure 2A, Table 1), this represents considerably fewer variants (n = 519, Table 2) than for other VUS subclasses. Interestingly, the VUS-high subclass did not have the most movement for every contributing laboratory. For two labs, the most reclassifications were among variants starting as LB or VUS-low (Figure S3).

**Figure 4.**
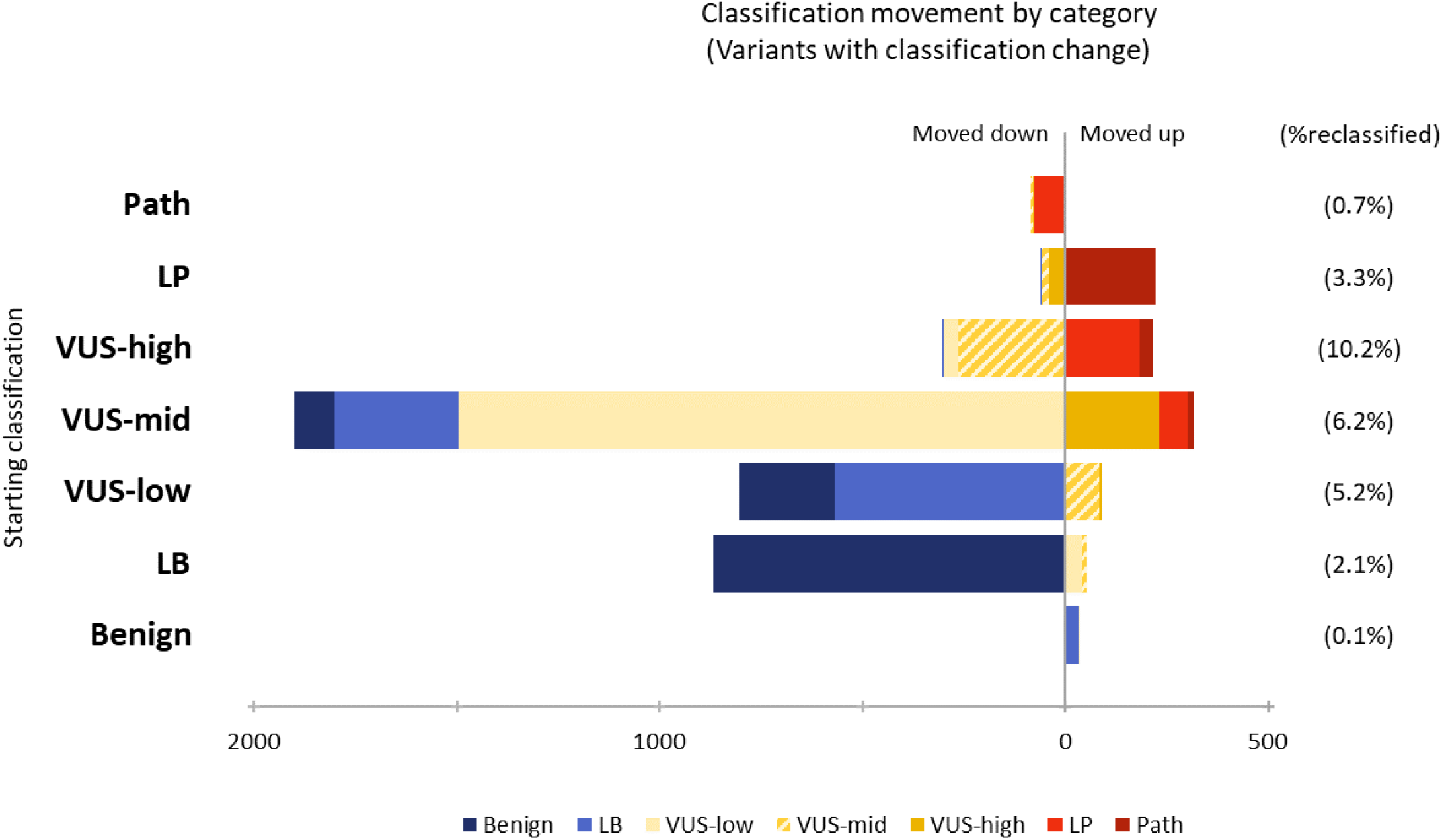
Variant classification movement including final classification. The figure shows the number of variants from four laboratories that changed classification. Variants’ initial (starting) classification during the study period is shown at left. Variants whose classification moved “down” (toward benign) are shown on the left of the central vertical line. Variants whose classification moved “up” (toward pathogenic) are shown to the right of the central vertical line. Colored segments represent the variants’ classification at the end of the study period. The percentage of variants that were reclassified up or down is shown at right. Abbreviations as in Figure 2.

While patterns of VUS reclassifications were unique for each contributing lab (Figure S3), overall, variant classifications starting at any VUS subclass were more likely to move down over time, and the odds of this were distinct for each subclass (Table 2, Figure 3). A variant with a starting classification of VUS-high was only 1.4 times more likely to move downward than upward, which is significantly less than when only a single VUS class is considered (OR=2.9, p < 0.0001, Table 2). On the other hand, variants with a starting classification of VUS-mid or VUS-low had significantly higher odds of moving down (6.0:1 and 9.0:1, respectively) when compared to a single VUS class (OR= 0.67 and 0.45, p < 0.0001, respectively).

Overall, the classification of 96.7% of variants were unchanged during our study period (Figure 3). The classification of these variants remains static for one of two reasons: 1) the variant was not evaluated again during the study period, or 2) the variant was re-evaluated but there were insufficient data to change the classification. To distinguish between these causes, we looked at the data from two labs (Labcorp and Quest Diagnostics) where all classification events were tracked through the study periods.

We found in both cases the vast majority of variants were evaluated only once (Figure S4A,C). Among variants evaluated at least twice during the study period, we still see the majority of variant classifications do not change upon re-evaluation (87.7% and 74.8% for Labcorp and Quest, respectively, Figure S4B,D).

In addition to how many variants moved up or down in classification, we also evaluated how far those variant classifications moved. For instance, we noted whether a VUS-low was upgraded to a higher VUS subclass, or further, to likely pathogenic or pathogenic. Indeed, when being upgraded, most VUS-low only moved to VUS-mid and none moved to pathogenic or likely pathogenic (Table 2, Figure 4). Atypical reclassifications can be found in Table S2 and S3 and included five VUS-low variants that moved to VUS-high and two that were reclassified as risk alleles (discussed below). Similarly, only two variants moved from VUS-high down to likely benign and none to benign (Table S2). While no variants starting as benign or likely benign moved up past VUS-mid, three variants starting at likely pathogenic moved down as far as likely benign. These classification changes were due to new evidence that became available after initial assessment, including two whose functional data was reassessed by a ClinGen expert panel and one that was in linkage disequilibrium with a disease allele where the causative variant has since been identified (Table S2).

## Discussion

In this analysis of four laboratories that utilize VUS subclasses, we found informative trends in the likelihood that variants in each subclass would move up or down across the major categories of variant classification. While the methods of how each laboratory applied a subclass were not identical, the implications of the experience of each laboratory were similar. Most importantly, we found the category of VUS-low never moved to a Mendelian P/LP classification (Table 2, Figure 4). It’s worth noting there were four variants initially classified as benign, likely benign, or VUS-low that were later reclassified as a likely or established risk allele for a common disease trait (Table S3). For example, one variant that was VUS-low for a monogenic disorder (MODY) later became a likely risk allele for Type 2 Diabetes. However, these were extremely rare events. These data suggest it may be permissible to consider de-emphasizing the category of VUS-low on reports, such as shifting to a supplement or excluding them from reports altogether. This would be particularly useful in settings where there is a low prior probability of a positive result (e.g., most germline cancer testing). Indeed, for Baylor Genetics’ panel-based hereditary cancer tests, variants classified as VUS-low are not included in reports unless the laboratory is requested to do so for a specific patient by the ordering clinician. With types of testing where there is a high prior probability of a positive result, there will often be a strong correlation between the patient’s phenotype and the gene in which a variant is identified, such that application of the specific phenotype criterion (PP4)^14^ could elevate classification of a variant with otherwise very little evidence, and ensure it is still considered and included on the report. This highlights the importance of providing phenotype information to the testing laboratory, so it can appropriately prioritize variants for reporting.^15^ It should be noted that updates to the 2015 ACMG/AMP guidelines, that have been presented for community input in advance of finalization, suggest it is likely many variants currently falling into our VUS-low categories will become likely benign. In addition, a subset in VUS-mid may become VUS-low. However, even if this is true, we believe changes in how the future VUS-low category is managed may still be warranted.

We found the VUS-high category of variants had the fewest number of variants but was the class most likely to change, with an overall rate of 10.2% change (Figure 4). These variants were also more likely to move up in classification (to likely pathogenic or pathogenic) than other VUS subclasses. These findings suggest laboratories, physicians, and patients should invest additional resources in building evidence for these variants, such as segregation analysis, deeper clinical evaluations, splicing studies, and even collaborations with research laboratories who could perform functional and other multi-omic studies. The distinct attributes of VUS-high also indicates these variants could be considered for reporting in certain population screening settings where there is better support for managing VUS and the basis and implications of uncertainty are better understood. Indeed, at MGB LMM, VUS-high (labeled as VUS-Favor Pathogenic) were reported in the MedSeq study^7^ with systems in place to record return of results sessions, have follow-up sessions with physicians and patients if questions arose, and intervene if mismanagement occurred. Separately, participants in the Illumina Understand Your Genome (UYG) received this subclass as “suspicious” VUS with educational sessions offered for participants to understand their results and ask questions.

The distinct rates and directions of reclassification observed across the VUS subclasses emphasizes the importance of clear communication of variant information by testing laboratories to healthcare providers and patients. This is particularly critical as the gene content for multi-gene panels grows due to the broadening of test indications, thereby increasing the VUS rate and decreasing the likelihood these VUS are relevant to the patient’s phenotype^1^. Communication of VUS subclasses, whether with specific terms (e.g., labeled as VUS-low, mid, or high) or how they are included/excluded or emphasized/deemphasized on clinical reports (e.g., using supplements or distinct sections of reports), can provide additional information about the potential clinical significance of a VUS, and therefore help providers make more informed decisions and offer clearer genetic counseling for patients and families. For VUS-high variants, patients could be instructed to participate in follow-up studies, share their data through patient registries, and track their variant over time for classification changes.

It should be noted that the experience of these four laboratories is based on laboratory-specific rules for variant subclasses that were likely variable given the absence of guidelines, variation in the genes and associated diseases being tested, policy for review of previous classifications, and how likely benign variants were managed (e.g., Baylor Genetics included only cancer genes and did not capture many likely benign variants, MGB LMM had higher rates of cardiovascular and hearing loss genes). These differences may account for the distinct ratios of current classifications as well as variability in variant reclassification by class for each laboratory (Figures S1 and S3). We anticipate that future versions of the ACMG/AMP variant classification standards, which will provide more concrete guidance on how VUS subclasses are applied, will enable more consistency across laboratories. This type of analysis can then be re-examined to further inform future use of VUS subclasses in laboratory reporting.

It is also important to note that for 96.7% of variants no classification change occurred (Figure 3), largely because variants were observed only a single time in the study period and therefore not subjected to re-analysis (Figure S4). This emphasizes the importance of sharing classifications and evidence broadly through databases like ClinVar and encouraging physicians and patients to utilize a laboratory’s re-analysis and variant clarification services. ClinVar intends to support the use of VUS subclasses once the new professional guidelines are published, if not before. Additionally, robust methods to connect clinical laboratory testing to research efforts are needed, so detailed phenotypes can be collected and additional studies into variant impact can be performed even if the variant is only observed a single time. It is through these community efforts that we anticipate making significant advances in understanding variants currently classified as uncertain significance.

In summary, with the high rate of VUS on clinical genetic testing reports for symptomatic testing, particularly MGP reports,^1^ there is a need to carefully consider how and when VUS are reported. There is currently limited guidance from professional organizations around VUS reporting except for carrier screening^9,10^ and currently no guidance around the reporting of VUS subclasses. Excessive VUS reporting can lead to negative impacts in terms of resource usage and improper clinical management, yet removing VUS from reports could prevent patients and clinicians from fully utilizing key services such as the laboratory’s reclassification or reanalysis services. We believe that utilization of VUS subclasses and the development of subclass-specific professional guidance is critically needed to guide our community in consistent practices optimized for improved patient outcomes and resource utilization.

## Data Availability

All four laboratories submit variant classifications to ClinVar. Laboratory-specific data used to inform this study can be found in Supplementary Tables S4-S11.

## Acknowledgements

The authors are thankful to the variant analysts and other staff at their respective laboratories for variant classification, infrastructural, scientific, and due diligence support.

## Author Contributions

Conceptualization: S.H., I.K., H.R.; Data curation: G.B., W.C., M.L., L.M., N.N, R.R.; Formal analysis: G.B., W.C., M.L., L.M., N.N.; Methodology: G.B., W.C., M.L., N.N.; Visualization: G.B.; Writing-original draft: G.B., I.K., H.R.; Writing-review & editing: G.B., I.K., M.L., L.M., N.N., H.R., R.R., S.H.

## Ethics Declaration

This study was not considered human subjects research given that no individual-level data was accessed.

## References

1. Rehm HL, Alaimo JT, Aradhya S, et al. The landscape of reported VUS in multi-gene panel and genomic testing: Time for a change. Genet Med. 2023;25(12):100947. doi:10.1016/j.gim.2023.100947

2. Richards S, Aziz N, Bale S, et al. Standards and guidelines for the interpretation of sequence variants: a joint consensus recommendation of the American College of Medical Genetics and Genomics and the Association for Molecular Pathology. Genet Med. 2015;17(5):405–423. doi:10.1038/gim.2015.30

3. Tavtigian SV, Greenblatt MS, Harrison SM, et al. Modeling the ACMG/AMP variant classification guidelines as a Bayesian classification framework. Genet Med. 2018;20(9):1054–1060. doi:10.1038/gim.2017.210

4. Tavtigian SV, Harrison SM, Boucher KM, et al. Fitting a naturally scaled point system to the ACMG/AMP variant classification guidelines. Hum Mutat. 2020;41(10):1734–1737. doi:10.1002/humu.24088

5. Niehaus A, Azzariti DR, Harrison SM, et al. A survey assessing adoption of the ACMG-AMP guidelines for interpreting sequence variants and identification of areas for continued improvement. Genet Med. 2019;21(8):1699–1701. doi:10.1038/s41436-018-0432-7

6. Karbassi I, Maston GA, Love A, et al. A Standardized DNA Variant Scoring System for Pathogenicity Assessments in Mendelian Disorders. Hum Mutat. 2015;37(1):127–134. doi:10.1002/humu.22918

7. McLaughlin HM, Ceyhan-Birsoy O, Christensen KD, et al. A systematic approach to the reporting of medically relevant findings from whole genome sequencing. BMC Méd Genet. 2014;15(1):134. doi:10.1186/s12881-014-0134-1

8. Durkie M, Cassidy EJ, Berry I, et al. ACGS Best Practice Guidelines. ACGS Best Practice Guidelines for Variant Classification in Rare Disease 2024. Updated February 20, 2024. Accessed March 8, 2024. https://www.acgs.uk.com/quality/best-practice-guidelines/#VariantGuideline

9. Deignan JL, Astbury C, Cutting GR, et al. CFTR variant testing: a technical standard of the American College of Medical Genetics and Genomics (ACMG). Genet Med. 2020;22(8):1288–1295. doi:10.1038/s41436-020-0822-5

10. Gregg AR, Aarabi M, Klugman S, et al. Screening for autosomal recessive and X-linked conditions during pregnancy and preconception: a practice resource of the American College of Medical Genetics and Genomics (ACMG). Genet Med. 2021;23(10):1793–1806. doi:10.1038/s41436-021-01203-z

11. Azzariti D, Biesecker LG, Harrison S. ClinGen Sequence Variant Interpretation. Updated December 12, 2023. Accessed March 8, 2024. https://clinicalgenome.org/working-groups/sequence-variant-interpretation/#heading_membership

12. Miller DT, Lee K, Gordon AS, et al. Recommendations for reporting of secondary findings in clinical exome and genome sequencing, 2021 update: a policy statement of the American College of Medical Genetics and Genomics (ACMG). Genet Med. 2021;23(8):1391–1398. doi:10.1038/s41436-021-01171-4

13. MedCalc Software Ltd. Odds ratio calculator. Version 22.032. Accessed July 23, 2024. https://www.medcalc.org/calc/odds_ratio.php

14. Biesecker LG, Byrne AB, Harrison SM, et al. ClinGen guidance for use of the PP1/BS4 co-segregation and PP4 phenotype specificity criteria for sequence variant pathogenicity classification. Am J Hum Genet. 2024;111(1):24–38. doi:10.1016/j.ajhg.2023.11.009

15. Bush LW, Beck AE, Biesecker LG, et al. Professional responsibilities regarding the provision, publication, and dissemination of patient phenotypes in the context of clinical genetic and genomic testing: points to consider—a statement of the American College of Medical Genetics and Genomics (ACMG). Genet Med. 2018;20(2):169–171. doi:10.1038/gim.2017.242

