## Supplement for "Distinct rates of VUS reclassification are observed when subclassifying VUS by evidence level"

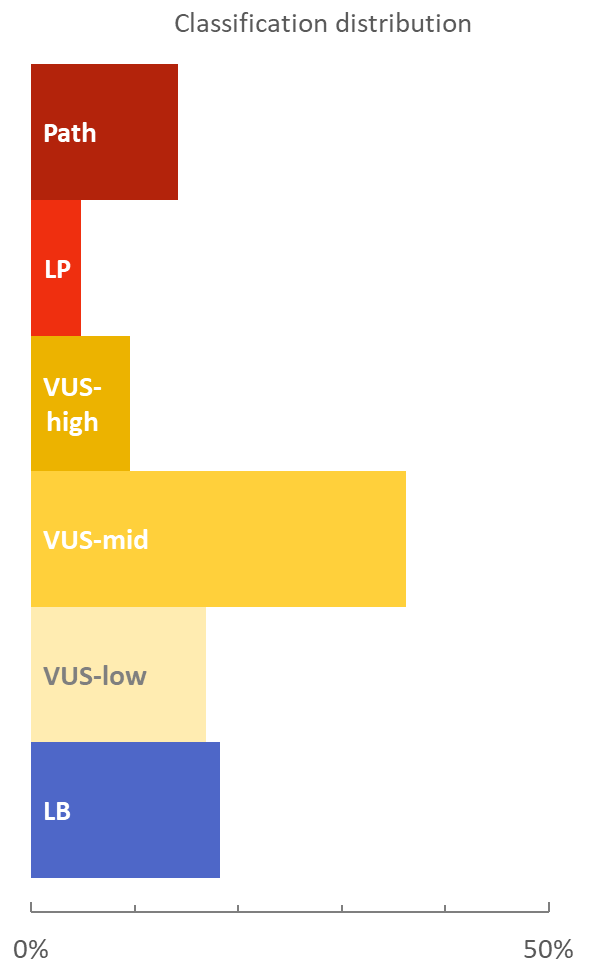

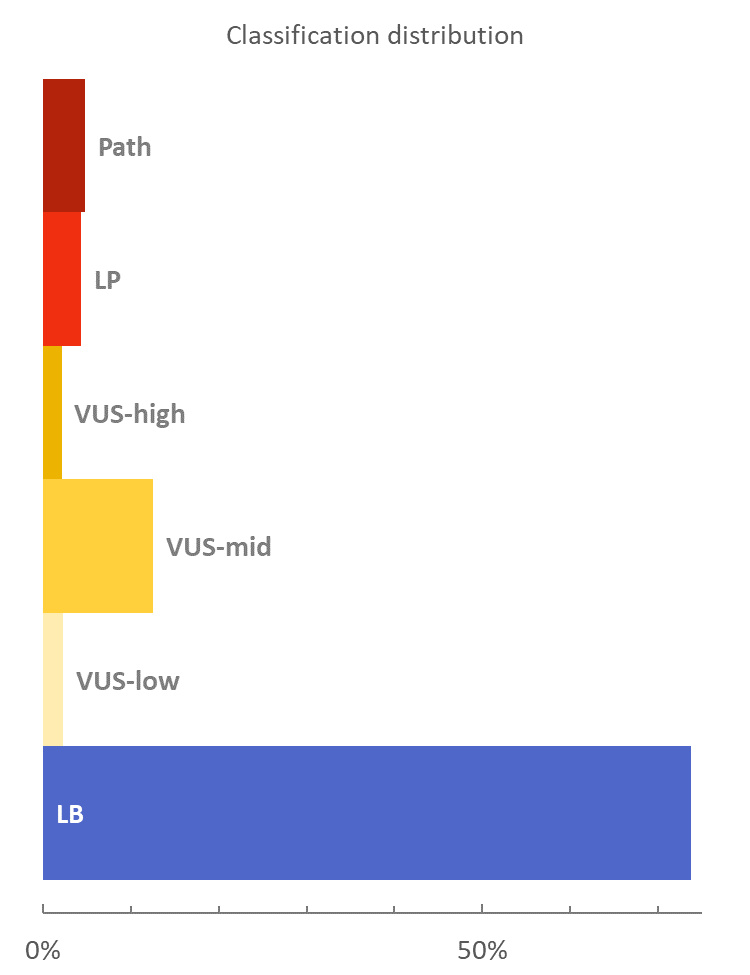

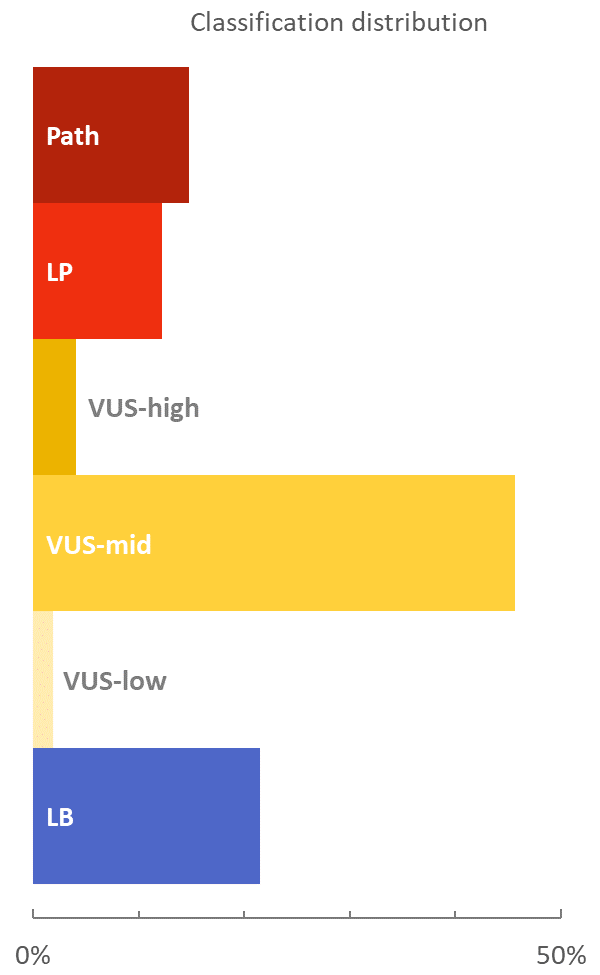

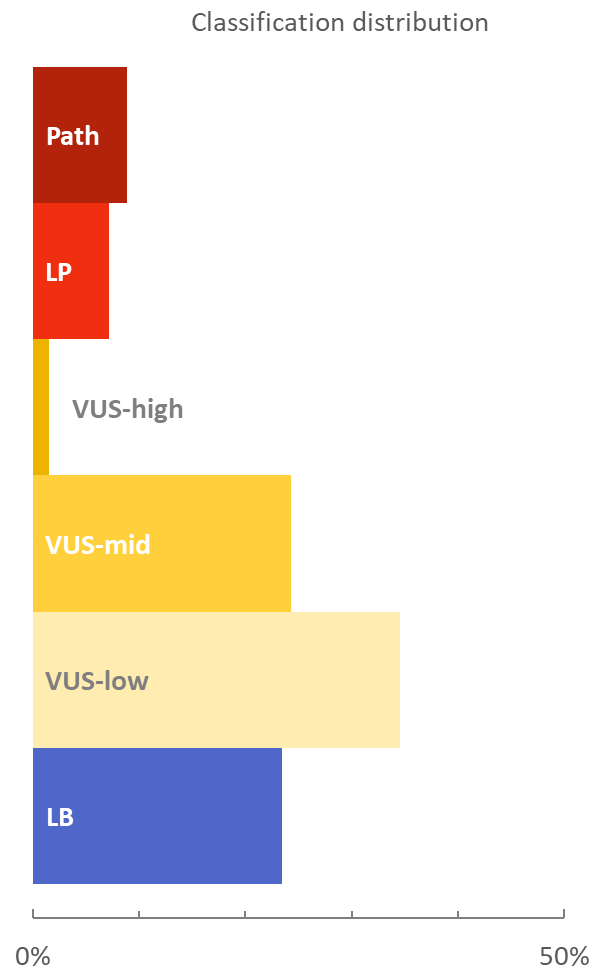
 A) Baylor Genetics	B) Labcorp	C) MGB LMM	D) Quest Diagnostics

### Figure S1. Variant classification distributions by laboratory.

As in Figure 2A, the distribution of variants across six variant classifications from likely benign to pathogenic, including three VUS subclasses are shown. Each panel displays data from a single laboratory, as indicated. This figure highlights differences in the percentage of variants that were classified across 6 classes/subclasses of variants. Differences are likely due to a combination of factors, such as: evidence thresholds for classification, the time between initial testing and any subsequent review, the timeframe for this study, distinct sets of genes tested in each lab, and attributes of those genes that contribute to disease. MGB LMM, Mass General Brigham Laboratory for Molecular Medicine; Path, pathogenic; LP, likely pathogenic; VUS, variant of uncertain significance; LB, likely benign.

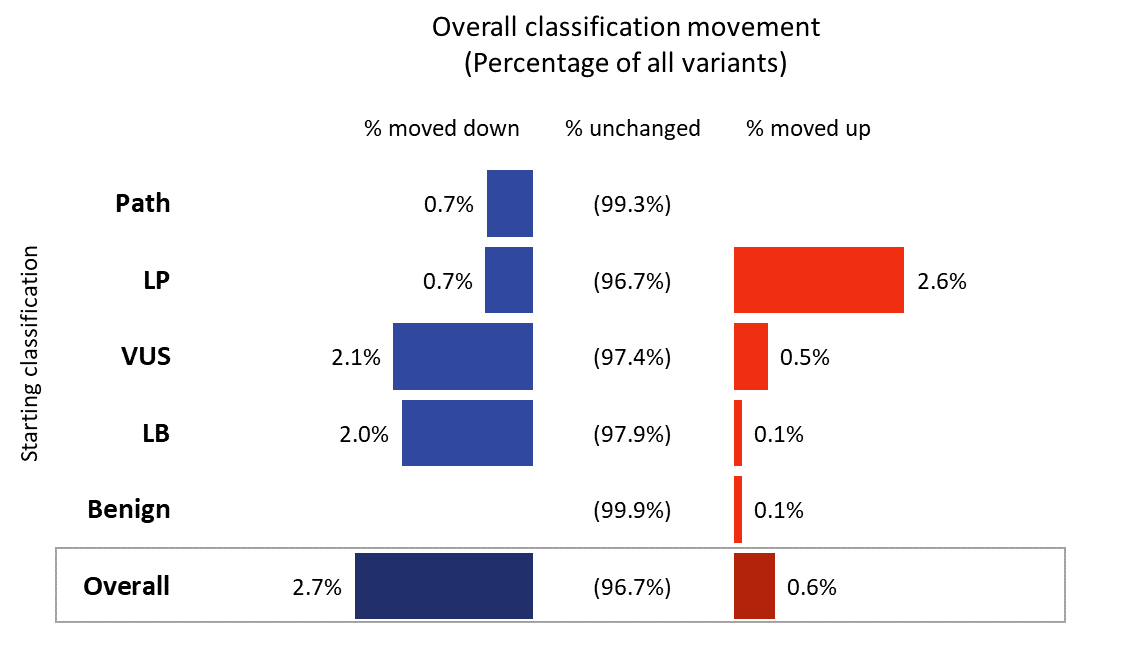

### Figure S2. Classification movement with a single VUS category.

As in Figure 3, the percentage of variants from the four laboratories that did or did not change classification, except VUS is shown as one category, rather than three subclasses. Variants’ initial (starting) classification during the study period is shown at left. Blue bars, the percentage of variants whose classification moved at least one category “down” (toward benign). Red bars, the percentage of variants whose classification moved at least one category “up” (toward pathogenic). Variants that were not re-evaluated or whose classification did not change during the study period are shown as “unchanged.” Path, pathogenic; LP, likely pathogenic; VUS, variant of uncertain significance; LB, likely benign.

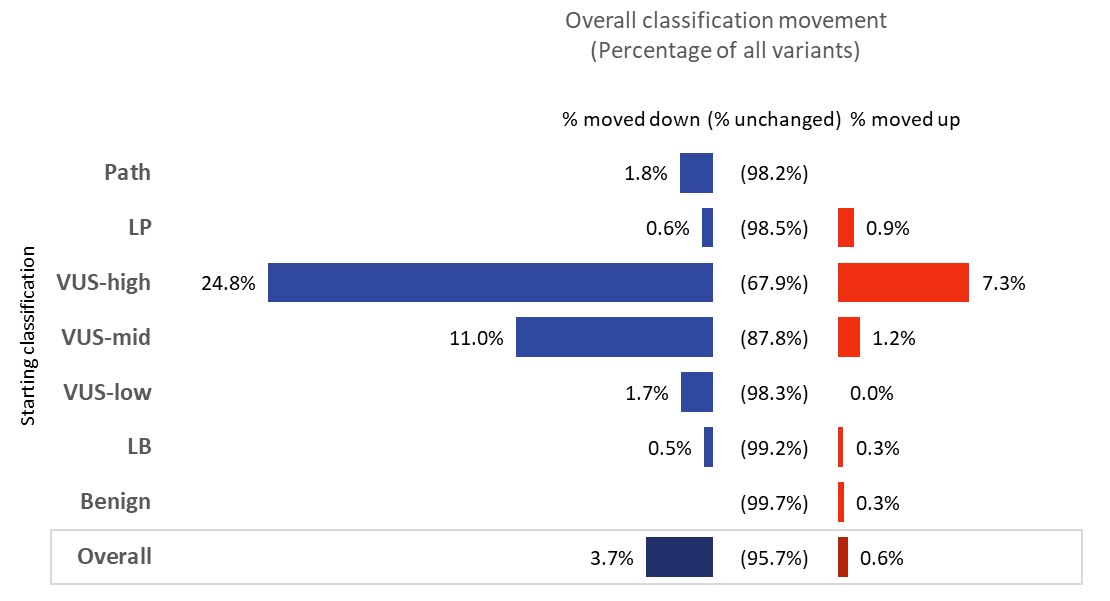

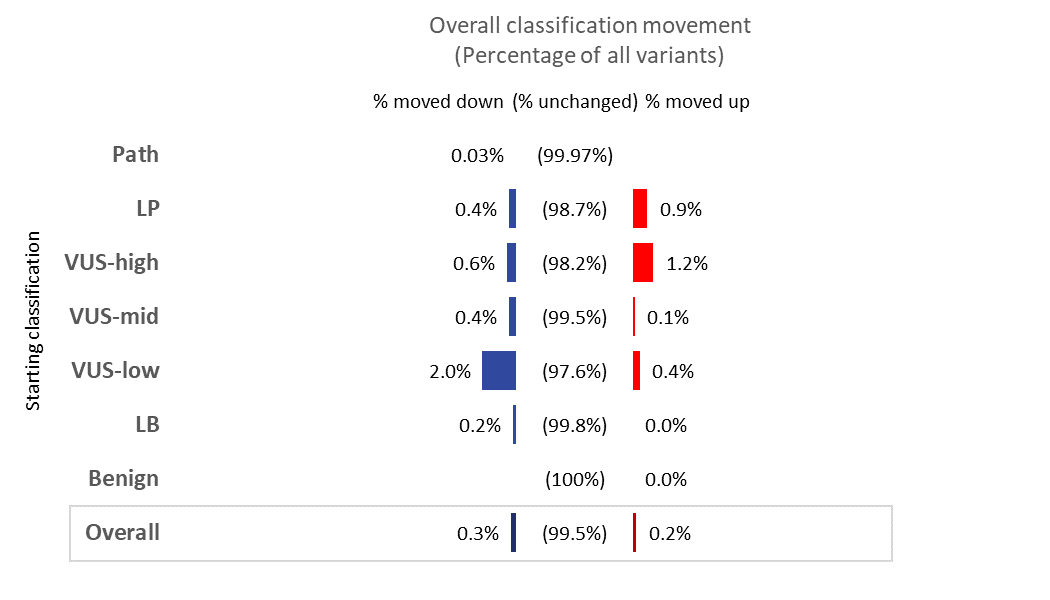
A) Baylor Genetics	B) Labcorp

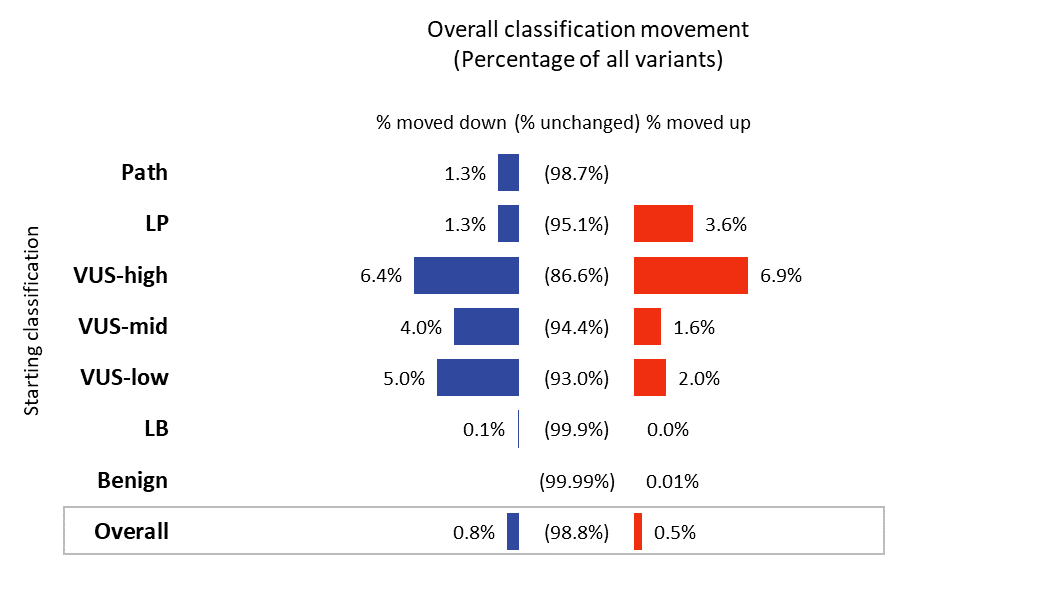

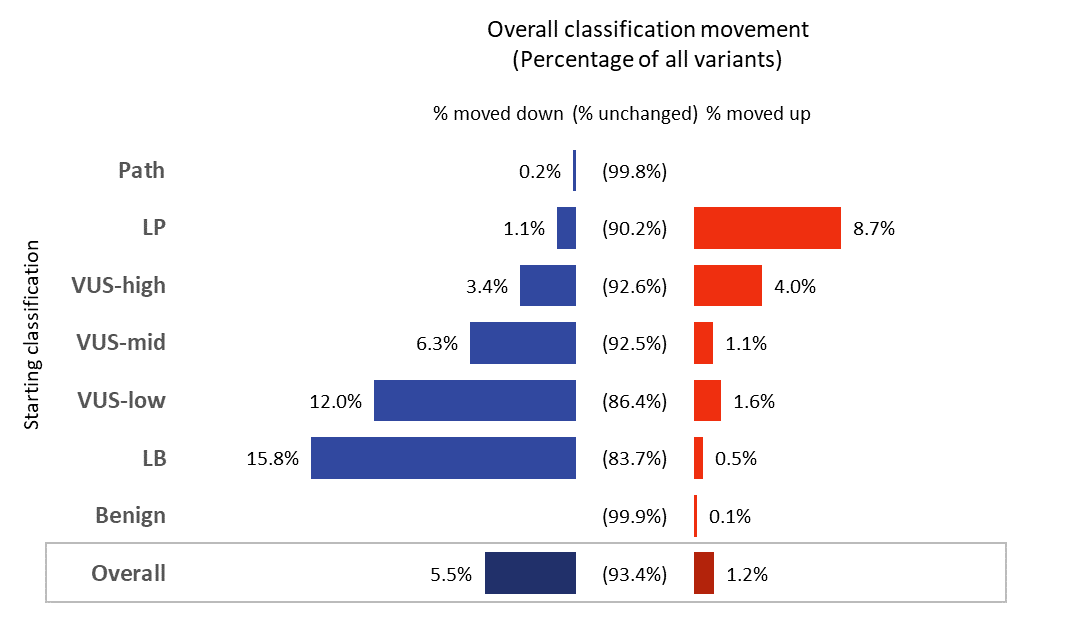
C) MGB LMM	D) Quest Diagnostics

### Figure S3. Classification movement by laboratory.

Percentage of variants that did or did not change classification for each of the four laboratories, as indicated. Variants’ initial (starting) classification during the study period is shown at left. Blue bars, the percentage of variants whose classification moved at least one category “down” (toward benign). Red bars, the percentage of variants whose classification moved at least one category “up” (toward pathogenic). Variants that were not re-evaluated or whose classification did not change during the study period are shown as “unchanged.” This figure highlights differences in the percentage of variants that were reclassified. Differences are likely due to a combination of factors, such as: evidence thresholds for classification, the time between initial testing and any subsequent review, the timeframe for this study, distinct sets of genes tested in each lab, and attributes of those genes that contribute to disease. MGB LMM, Mass General Brigham Laboratory for Molecular Medicine; Path, pathogenic; LP, likely pathogenic; VUS, variant of uncertain significance; LB, likely benign.

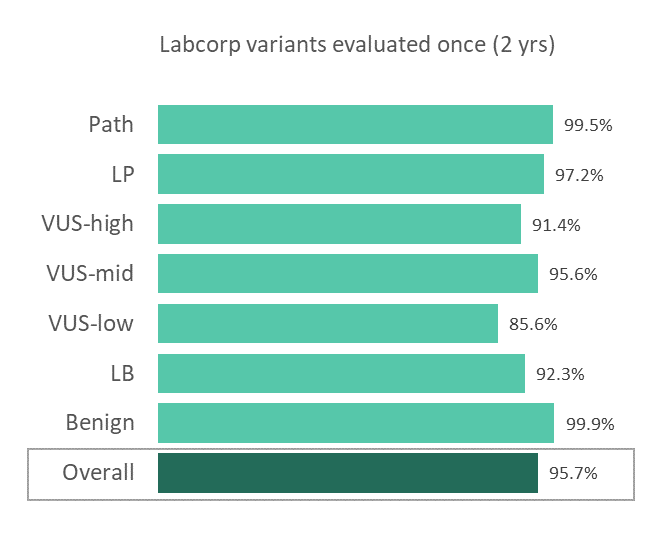

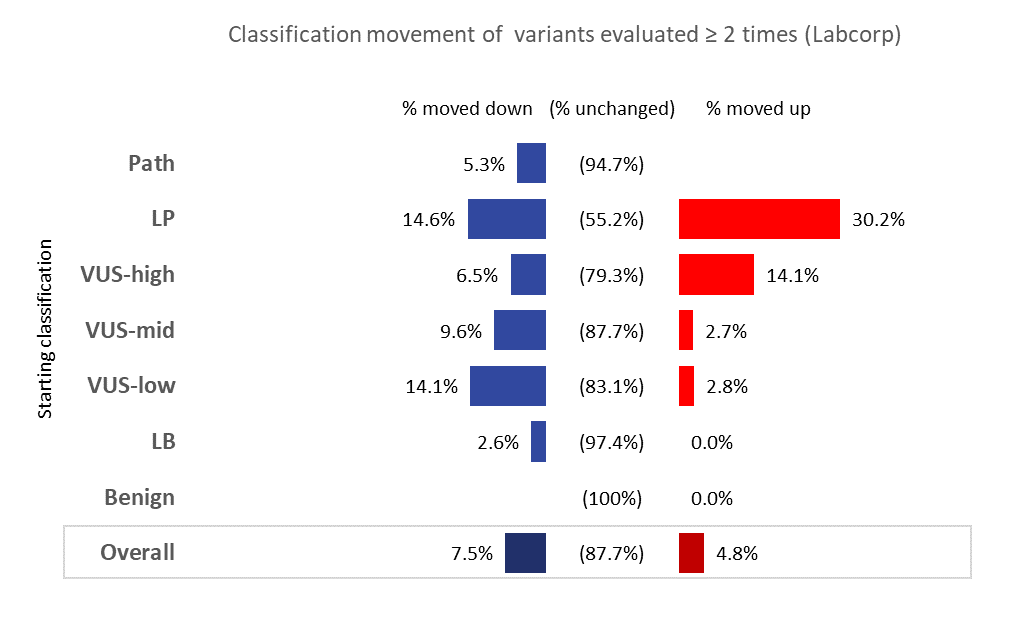
A)	B)

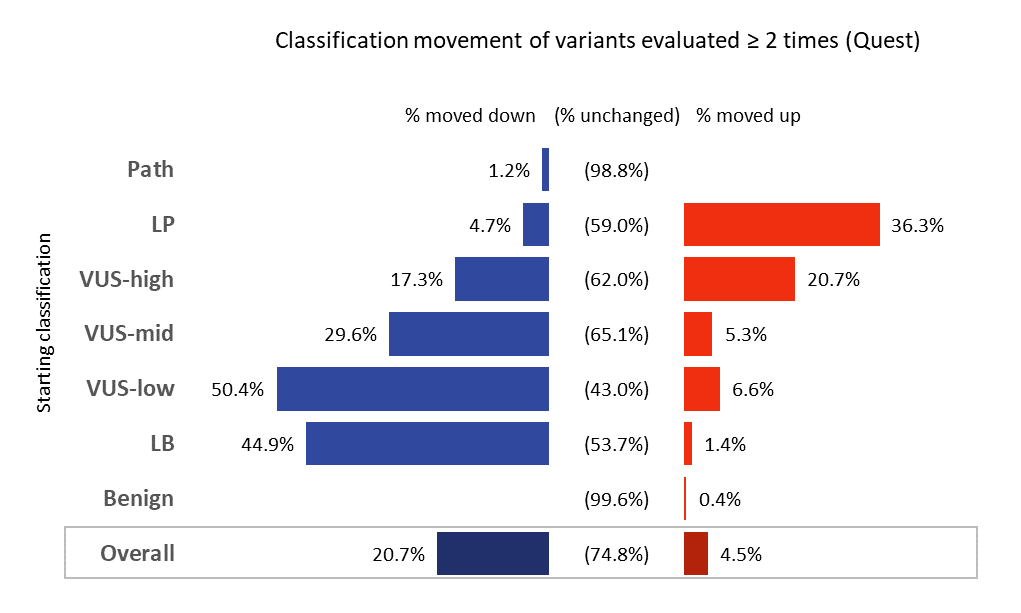

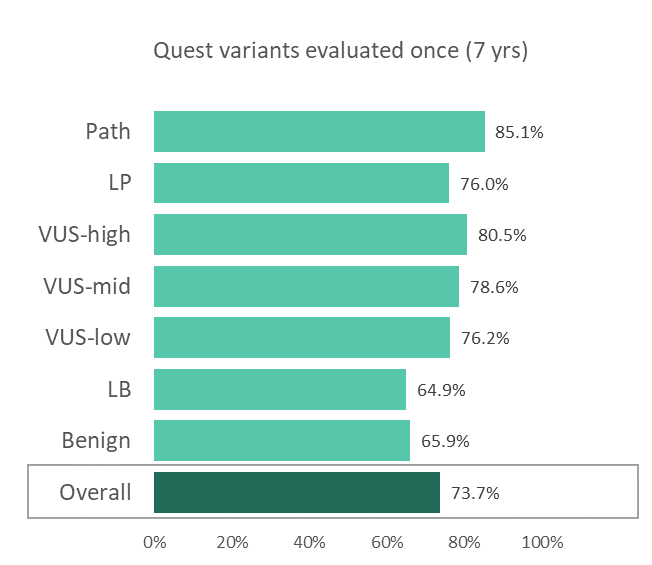
C)	D)

### Figure S4. Classification movement stratified by re-evaluation status.

(A,C) Percentage of variants in each class that were evaluated a single time during the study period for two of the contributing laboratories. (B,D) The percentage of variants that did or did not change classification, as described in Figure S4, except variants evaluated once during the study period have been removed. Path, pathogenic; LP, likely pathogenic; VUS, variant of uncertain significance; LB, likely benign.

### Table S1. Indication of VUS subclass on clinical reports, by laboratory.

|  | **VUS-low** | **VUS-mid** | **VUS-high** |
| --- | --- | --- | --- |
| **Baylor Genetics** | [Internal use only; not typically reported] | [Internal use only; reported as VUS] | [Internal use only; reported as VUS] |
| **Labcorp** | [Primarily for internal use; when reportable and distinguished, language is as follows:]  *GENE* c.XXX (p.XXX) (VUS - possibly benign). Evidence suggests that the variant may be benign, although additional information is needed. | *GENE*  c.XXX (p.XXX) (VUS - uncertain significance).  Insufficient information is available to classify this variant as either benign or pathogenic. | [Primarily for internal use and reported as VUS; when distinguished, language is as follows:]  *GENE*  c.XXX (p.XXX) (VUS - possibly pathogenic). Evidence suggests that the variant may be pathogenic, although additional information is needed. |
| **Mass General Brigham Laboratory for Molecular Medicine** | In summary, while the clinical significance of this variant is uncertain, these data/its frequency suggests that it is more likely to be benign. | In summary, the clinical significance of this variant is uncertain | In summary, while there is some suspicion for a pathogenic role, the clinical significance of this variant is uncertain. |
| **Quest Diagnostics, Neurology (Athena Diagnostics)** | “*GENE* c.XXX (p.XXX) is classified as a variant of uncertain significance (VUS). While the following *data point to a lack of association with disease*, there is insufficient evidence to determine the clinical significance of the variant at this time:”  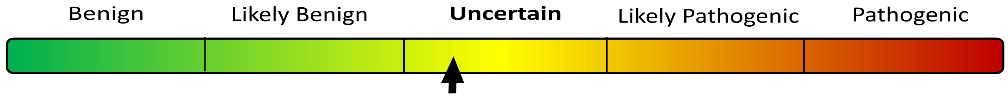 | “*GENE* c.XXX (p.XXX) is classified as a variant of uncertain significance (VUS). The following data are insufficient to determine the clinical significance of the variant at this time:”  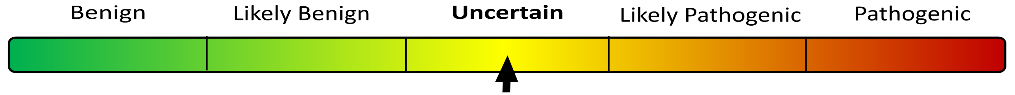 | “*GENE* c.XXX (p.XXX) is classified as a variant of uncertain significance (VUS). While the following *data point to a possible association with disease*, there is insufficient evidence to determine the clinical significance of the variant at this time:”  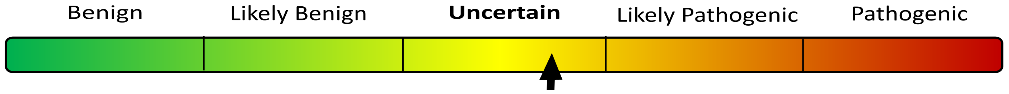 |

### Table S2. Variants with atypical classification shifts during the study period.

| Variant | Gene symbol (ID) | Starting class | Ending class | Comments |
| --- | --- | --- | --- | --- |
| NC_000010.10:g.89623056C>T (NM_000314.4:c.-1171C>T) | PTEN (HGNC:9588) | LP | LB | Experimental data indicating a reduction in promoter activity for the PTEN gene, together with a lack of population frequency, supported pathogenicity. Later assessment of the experimental results, informed by the ClinGen PTEN Expert panel, considered them to be insufficient to support pathogenicity. Additional data provided by members of the expert panel indicating a lack of disease association, together with updated population data, led to a likely benign classification. |
| NC_000010.10:g.89623031_89623042del  (NM_000314.4:c.-1196_-1185del) | PTEN (HGNC:9588) | LP | LB | Experimental data indicating a reduction in promoter activity for the PTEN gene, together with low population frequency, supported pathogenicity. Re-evaluation of the population data moved classification to uncertain. Later assessment of the experimental results, informed by the ClinGen PTEN Expert panel, considered them to be insufficient to support pathogenicity. Additional data provided by members of the expert panel indicated a lack of disease association and led to a likely benign classification. |
| NM_000256.3:c.3628-41_3628-17del | MYBPC3 (HGNC:7551) | LP | LB | Variant initially considered likely pathogenic for hypertrophic cardiomyopathy with low penetrance in the heterozygous state and increased penetrance in the homozygous state. Subsequent literature established a deep intronic variant as the causative allele and this variant as a "tag" identifiable by panel and exome sequencing. |
| NM_133261.3:c.787+5G>C | GIPC3 (HGNC:18183) | VUS-high | LB | Initially identified homozygous in affected proband and siblings, and population data was not available to refute a possible disease association. Family testing identified the variant homozygous in unaffected parents, indicating lack of segregation with disease. |
| NM_000179.2:c.3801+5G>A | MSH6 (HGNC:7329) | VUS-high | LB | In silico algorithms predict a negative impact on RNA splicing, however, later studies showed no abnormal splicing to be detected. Re-evaluation of population data, together with new clinical evidence, indicated a lack of association with disease. |
| NM_000527.5:c.1078G>C | LDLR (HGNC:6547) | VUS-low | VUS-high | Variant was initially called a VUS-low due to limited case data and a high frequency in the Latino population (0.11%). Subsequent reanalysis identified a new paper suggesting the variant is a founder variant in the Mexican population. |
| NM_033380.3:c.646-6C>G | COL4A5 (HGNC:2207) | VUS-low | VUS-high | Computational predictions changed to suggest a possible impact on RNA splicing. |
| NM_000435.2:c.6102dup | NOTCH3 (HGNC:7883) | VUS-low | VUS-high | Initial assessment of population data suggested benignity. Subsequent updates to population data no longer supported that assessment. |
| NM_000492.3:c.4056G>C | CFTR (HGNC:1884) | VUS-low | VUS-high* | Impact of available population data on the classification changed due to internal criteria.  *Since the end of the study period, this variant’s classification has shifted back to VUS-low. |
| NM_000162.3:c.680-15C>A | GCK (HGNC:4195) | VUS-low | VUS-high | Computational predictions changed to suggest a possible impact on RNA splicing. |
| NM_153638.4:c.1133A>G | PANK2 (HGNC:15894) | VUS-low | VUS-high | Initial - Variant with higher frequency in East Asian subpopulations above our threshold for recessive disease. Case counts reported with other VUS compound heterozygote genotypes were conservatively ascertained resulting in a VUS-low. Re-evaluation - A new patient carrying this variant and a likely pathogenic variant reported and re-evaluation of previous ascertainment to capture additional compound heterozygous case counts led to the VUS-high categorization. At-least one additional submitter re-classified the variant from "Likely benign" to "Pathogenic" since our initial evaluation. |
| LP, Likely pathogenic; LB, Likely benign; VUS, variant of uncertain significance. | | | | |

### Table S3. Variants reclassified as risk allele during the study period.

| Variant | Gene symbol (ID) | Starting classification | Ending classification | Comments |
| --- | --- | --- | --- | --- |
| NM_000545.5:c.1522G>A | HNF1A (HGNC:11621) | VUS-low | Likely  Risk allele | Gene is included on a panel for monogenic diabetes of the young (MODY), and evidence for this variant was not supportive of pathogenicity for that test indication. Later evidence supports this variant as a likely risk allele for type 2 diabetes instead. |
| NM_000248.4:c.952G>A | MITF (HGNC:7105) | VUS-low | Established  Risk allele | Gene was initially tested as part of a hearing loss panel, where its high frequency is too common for rare genetic hearing loss and Oculocutaneous albinism. Subsequent analyses for genomic screening evaluated case control studies detailing evidence for this variant as a risk allele for melanoma. |
| NM_000219.6:c.253G>A | KCNE1 (HGNC:6240) | LB | Likely  Risk allele | Gene was initially tested as part of a hearing loss panel, where its high frequency is too common for Jervell and Lange-Nielsen syndrome. Subsequent analyses for genomic screening evaluated case control studies detailing evidence for this variant as a risk allele for long QT syndrome. |
| NM_025225.3:c.444C>G | PNPLA3 (HGNC:18590) | Benign | Established  Risk allele | Variant was initially identified in a genomic screen of high penetrance disorders and classified as benign based upon very high population frequency. A subsequent focus on adding risk alleles to genomic screening programs identified this variant with an association to non-fatty acid liver disease in the homozygous state. |
| LB, Likely benign; VUS, variant of uncertain significance. | | | | |

### Table S4. Variant classification distribution: Baylor Genetics

|  | **Variant Type** | | | | | | | | | |  | |
| --- | --- | --- | --- | --- | --- | --- | --- | --- | --- | --- | --- | --- |
| **Classification** | Canonical splice site | Frameshift | Nonsense | Small in-frame ins/del | Missense | Synony-mous | Intronic | Other non-coding (e.g., UTR) | Start-loss | Stop-loss | | **Total** |
| Pathogenic | 378 | 1206 | 1002 | 36 | 407 | 9 | 56 | 3 | 33 | - | | **3130** |
| LP | 502 | 992 | 569 | 24 | 330 | 10 | 75 | 4 | 37 | - | | **2543** |
| VUS-high | 12 | 16 | 19 | 6 | 441 | 6 | 37 | 6 | 3 | - | | **546** |
| VUS-mid | 11 | 37 | 28 | 208 | 8013 | 22 | 198 | 42 | 13 | - | | **8572** |
| VUS-low | 8 | 11 | 23 | 103 | 5350 | 1489 | 4084 | 1128 | 2 | - | | **12198** |
| LB | 1 | 9 | 6 | 31 | 1904 | 2693 | 3344 | 302 | 0 | - | | **8290** |

### Table S5. Variant classification distribution: Labcorp

|  | **Variant Type** | | | | | | | | | |  | |
| --- | --- | --- | --- | --- | --- | --- | --- | --- | --- | --- | --- | --- |
| **Classification** | Canonical splice site | Frameshift | Nonsense | Small in-frame ins/del | Missense | Synony-mous | Intronic | Other non-coding (e.g., UTR) | Start-loss | Stop-loss | | **Total** |
| Pathogenic | 369 | 983 | 763 | 67 | 1475 | 1 | 42 | 25 | 17 | 2 | | **3744** |
| LP | 675 | 773 | 462 | 46 | 1043 | 0 | 24 | 34 | 51 | 2 | | **3110** |
| VUS-high | 32 | 25 | 14 | 26 | 870 | 1 | 38 | 18 | 6 | 2 | | **1032** |
| VUS-mid | 138 | 264 | 174 | 254 | 9026 | 19 | 1353 | 289 | 49 | 23 | | **11589** |
| VUS-low | 3 | 1 | 0 | 7 | 378 | 0 | 83 | 10 | 0 | 0 | | **482** |
| LB | 6 | 4 | 5 | 40 | 2229 | 2399 | 743 | 23 | 1 | 1 | | **5451** |

### Table S6. Variant classification distribution: Mass General Brigham Laboratory for Molecular Medicine

|  | **Variant Type** | | | | | | | | | |  | |
| --- | --- | --- | --- | --- | --- | --- | --- | --- | --- | --- | --- | --- |
| **Classification** | Canonical splice site | Frameshift | Nonsense | Small in-frame ins/del | Missense | Synony-mous | Intronic | Other non-coding (e.g., UTR) | Start-loss | Stop-loss | | **Total** |
| Pathogenic | 167 | 428 | 420 | 21 | 533 | 10 | 50 | 6 | 6 | 1 | | **1642** |
| LP | 176 | 368 | 311 | 14 | 577 | 8 | 36 | 5 | 11 | 0 | | **1506** |
| VUS-high | 100 | 71 | 63 | 13 | 451 | 13 | 30 | 4 | 11 | 1 | | **757** |
| VUS-mid | 127 | 185 | 147 | 63 | 3478 | 28 | 235 | 57 | 25 | 9 | | **4354** |
| VUS-low | 7 | 8 | 13 | 6 | 691 | 20 | 51 | 12 | 4 | 0 | | **812** |
| LB | 5 | 13 | 11 | 53 | 2084 | 17872 | 5664 | 90 | 0 | 0 | | **25792** |

### Table S7. Variant classification distribution: Quest Diagnostics

|  | **Variant Type** | | | | | | | | | |  | |
| --- | --- | --- | --- | --- | --- | --- | --- | --- | --- | --- | --- | --- |
| **Classification** | Canonical splice site | Frameshift | Nonsense | Small in-frame ins/del | Missense | Synony-mous | Intronic | Other non-coding (e.g., UTR) | Start-loss | Stop-loss | | **Total** |
| Pathogenic | 261 | 1339 | 934 | 47 | 1045 | 9 | 173 | 31 | 43 | 7 | | **3889** |
| LP | 118 | 392 | 177 | 25 | 479 | 12 | 103 | 4 | 9 | 1 | | **1320** |
| VUS-high | 15 | 21 | 6 | 57 | 2276 | 25 | 198 | 8 | 4 | 1 | | **2611** |
| VUS-mid | 13 | 56 | 19 | 290 | 8502 | 410 | 313 | 282 | 5 | 8 | | **9898** |
| VUS-low | 4 | 6 | 4 | 42 | 2201 | 1577 | 734 | 54 | 6 | 1 | | **4629** |
| LB | 10 | 6 | 7 | 135 | 4313 | 9018 | 1957 | 217 | 5 | 1 | | **15669** |

### Table S8. Variant classification movement: Baylor Genetics

|  | **Ending classification** | | | | | | |
| --- | --- | --- | --- | --- | --- | --- | --- |
| **Starting**  **classification** | Benign | LB | VUS-  low | VUS-  mid | VUS-  high | LP | Path |
| Pathogenic | 0 | 0 | 0 | 4 | 0 | 53 | 3098 |
| LP | 0 | 0 | 2 | 3 | 10 | 2440 | 22 |
| VUS-high | 0 | 0 | 22 | 137 | 435 | 39 | 8 |
| VUS-mid | 0 | 0 | 1051 | 8428 | 101 | 12 | 5 |
| VUS-low | 17 | 180 | 11097 | 0 | 0 | 0 | 0 |
| LB | 41 | 8095 | 24 | 0 | 0 | 0 | 0 |
| Benign | 5486 | 16 | 2 | 0 | 0 | 0 | 0 |

### Table S9. Variant classification movement: Labcorp

|  | **Ending classification** | | | | | | |
| --- | --- | --- | --- | --- | --- | --- | --- |
| **Starting**  **classification** | Benign | LB | VUS-  low | VUS-  mid | VUS-  high | LP | Path |
| Pathogenic | 0 | 0 | 0 | 0 | 0 | 1 | 3972 |
| LP | 0 | 0 | 0 | 10 | 4 | 3336 | 29 |
| VUS-high | 0 | 0 | 0 | 6 | 1045 | 12 | 1 |
| VUS-mid | 2 | 38 | 10 | 11816 | 10 | 4 | 0 |
| VUS-low | 0 | 10 | 482 | 1 | 1 | 0 | 0 |
| LB | 11 | 5450 | 0 | 0 | 0 | 0 | 0 |
| Benign | 2160 | 0 | 0 | 0 | 0 | 0 | 0 |

### Table S10. Variant classification movement: Mass General Brigham Laboratory for Molecular Medicine

|  | **Ending classification** | | | | | | |
| --- | --- | --- | --- | --- | --- | --- | --- |
| **Starting**  **classification** | Benign | LB | VUS-  low | VUS-  mid | VUS-  high | LP | Path |
| Pathogenic | 0 | 0 | 1 | 2 | 2 | 17 | 1665 |
| LP | 0 | 1 | 0 | 3 | 16 | 1455 | 55 |
| VUS-high | 0 | 1 | 5 | 43 | 661 | 48 | 5 |
| VUS-mid | 7 | 54 | 114 | 4141 | 38 | 30 | 4 |
| VUS-low | 13 | 28 | 760 | 15 | 1 | 0 | 0 |
| LB | 31 | 25779 | 2 | 3 | 0 | 0 | 0 |
| Benign | 9106 | 1 | 0 | 0 | 0 | 0 | 0 |

### Table S11. Variant classification movement: Quest Diagnostics

|  | **Ending classification** | | | | | | |
| --- | --- | --- | --- | --- | --- | --- | --- |
| **Starting**  **classification** | Benign | LB | VUS-  low | VUS-  mid | VUS-  high | LP | Path |
| Pathogenic | 0 | 0 | 0 | 1 | 0 | 6 | 3882 |
| LP | 0 | 2 | 0 | 2 | 9 | 1190 | 115 |
| VUS-high | 0 | 1 | 8 | 79 | 2418 | 84 | 21 |
| VUS-mid | 91 | 213 | 322 | 9160 | 84 | 23 | 5 |
| VUS-low | 204 | 352 | 4000 | 67 | 4 | 0 | 0 |
| LB | 785 | 4173 | 14 | 11 | 0 | 0 | 0 |
| Benign | 10670 | 15 | 1 | 0 | 0 | 0 | 0 |
